# Optimizing Atlas Counts for MRI-Guided Atlas-Based Autosegmentation of Swallowing Muscles in Head and Neck Radiotherapy

**DOI:** 10.1101/2025.07.28.25331930

**Authors:** Zayne Belal, Kareem Abdul Wahid, Sonja Stieb, Rachel Drummey, Christina S. Sharafi, Katherine A. Hutcheson, Stephen Y. Lai, Clifton D. Fuller, Brigid McDonald

**Affiliations:** Department of Radiation Oncology, University of Pennsylvania, Philadelphia, PA, USA; Department of Radiation Oncology, The University of Texas MD Anderson Cancer Center, Houston, TX, USA; Department of Radiation Oncology, Kantonsspital Arau, Arau, Switzerland; Department of Medicine, Inova Fairfax Hospital, Falls Church, VA, USA; College of Osteopathic Medicine, NOVA Southeastern University, Fort Lauderdale, FL, US; Department of Head and Neck Surgery, Division of Surgery, The University of Texas MD Anderson Cancer Center, Houston, TX, USA

**Keywords:** Atlas-based autosegmentation, MRI, head and neck cancer, swallowing muscles, radiation therapy, segmentation accuracy

## Abstract

**Purpose:** Radiotherapy-induced dysphagia can significantly impair head and neck (H&N) cancer patients’ quality of life. Despite the dose-dependent relationship between radiotherapy and dysphagia, swallowing structures are not routinely contoured due to time and labor demands. We evaluated atlas-based autosegmentation (ABAS) on MRI, identifying the optimal number of atlases required to efficiently and accurately delineate swallowing structures.

**Methods:** This study included pre-radiotherapy simulation T2-weighted MRIs from 60 H&N cancer patients enrolled in an IRB-approved observational trial. Scans were acquired on a 1.5T Siemens Aera scanner with H&N immobilization. Swallowing structures, including epiglottis, constrictors, digastric muscles, genioglossus, and others, were manually contoured for 25 atlas patients and 35 test patients. GTV-involved structures were excluded. ABAS was performed with increasing numbers of atlases (1-25) using a random-forest algorithm (ABAS-ADMIRE; Elekta) to determine the optimal atlas count. To mitigate variability from atlas selection, bootstrap resampling was implemented. Dice similarity coefficient (DSC), surface DSC (SDSC), average surface distance (ASD), and 95% Hausdorff distance (HD95) were calculated for each structure. Median computation times were calculated for each atlas count. Hsu’s MCB analysis identified the minimum atlas number statistically equivalent to the best-performing atlas range.

**Results:** Across all structures and metrics, Hsu’s analysis demonstrated that 2-4 atlases performed similarly to the best-performing atlas count. All structures except constrictors achieved median DSC>0.75 with ≥2 atlases. Computation times increased linearly with atlas count (range: ∼22-950 seconds for 1-25 atlases). These findings highlight that smaller atlas counts achieve comparable accuracy while significantly improving time efficiency.

**Conclusion:** Atlas-based autosegmentation is useful for delineating swallowing muscles in radiotherapy, especially with limited available contoured datasets. Utilizing 2-4 atlases achieves similar geometric accuracy to larger atlas counts, significantly reducing computational time without compromising clinical quality. This balance between efficiency and accuracy supports integration into workflows for better dysphagia prediction and treatment planning.

## Introduction

Head and neck cancer (HNC) accounts for approximately 4% of all cancers in the United States. Among the many treatment-related toxicities, dysphagia is one of the most common and functionally debilitating complications following radiotherapy (RT) for HNC patients [1]. Radiation dose to specific swallowing-related structures, including the pharyngeal constrictors and supraglottic larynx, has been correlated with the development of late dysphagia [2–4]. However, these structures are rarely contoured in routine clinical practice, in part due to their small size, large number, and the significant time burden associated with manual contouring [5,6]. As a result, dose estimation and toxicity modeling for these critical organs-at-risk (OARs) remains limited.

Magnetic resonance imaging (MRI) offers superior soft tissue contrast compared to computed tomography (CT), enabling better visualization of fine muscular anatomy, including many swallowing structures that are difficult to delineate on CT [7]. Despite this advantage, most prior auto-segmentation research in HNC has focused on CT, with limited studies investigating MRI-based delineation [8–12]. The ability to segment swallowing muscles on MRI may facilitate more accurate dose estimation and support the development of predictive models of dysphagia, particularly as functional MRI biomarkers of swallowing toxicity continue to emerge.

Atlas-based auto-segmentation (ABAS) is a widely used approach for automated contouring of normal tissues and target volumes in RT planning. It propagates labeled regions-of-interest (ROIs) from a set of manually contoured atlases to a new patient scan using deformable image registration (DIR), followed by label fusion to generate a consensus segmentation [9,13,14]. Utilization of multiple atlases has been shown to reduce errors caused by anatomical variation between atlas and patient, improving segmentation accuracy and robustness [9,13,14]. Additionally, ABAS has been combined with machine learning methods such as random forest classifiers to enhance voxel-level classification accuracy [15,16]. When trained effectively, ABAS outputs can support the generation of normal tissue complication probability (NTCP) models and enable large-scale retrospective and prospective analyses [17].

To date, no studies have systematically evaluated atlas-based auto-segmentation of swallowing-related muscles on MRI in HNC patients. Moreover, the number of atlases required to achieve optimal segmentation accuracy on T2-weighted MRI remains undefined. In this study, we investigate the performance of multi-atlas ABAS for segmenting swallowing-related muscles on T2-weighted MRI in patients with HNC and aim to quantify the minimum the minimum number of atlas patients necessary for accurate, time-efficient segmentation suitable for clinical and research applications.

## Methods

### Patients and Informed Consent

This study was a secondary analysis on a subset of patients enrolled in a prospective observational trial cohort (NCT03145077) conducted at MD Anderson Cancer Center (IRB approval number: PA16-0302). All patients gave written informed consent to participate in the study and for their images to be used.

### Imaging and Image Segmentation

Pre-RT simulation images were acquired in an RT immobilization mask, and T2-weighted MRIs were obtained on a 1.5 T Siemens Aera scanner with the following imaging parameters: 2-D turbo spin echo, repetition time = 4800 ms, echo time = 80 ms, slice thickness = 2 mm, pixel size = 0.5 mm. Inclusion criteria included adult patients with HNC who were planned to undergo RT, did not have any prior RT, and did not have osteoradionecrosis. Scans from 60 patients were included in this study. Table 1 shows the demographic information of the patient population.

**Table 1.** Demographic information of the patient population.

| Category | Subcategory | Count |
| --- | --- | --- |
| Age at Imaging | 18-45 (years) | 5 |
|  | 46-64 | 32 |
|  | 65+ | 23 |
| Gender | Female | 7 |
|  | Male | 53 |
| Race | American Indian or Alaska Native | 1 |
|  | Black or African American | 1 |
|  | Other | 2 |
|  | White or Caucasian | 56 |
| Smoking Status | Current | 5 |
|  | Former | 24 |
|  | Never | 31 |
| AJCC 7 Stage | N/A | 7 |
|  | Stage I | 13 |
|  | Stage II | 5 |
|  | Stage III | 15 |
|  | Stage IVA | 19 |
|  | Stage IVB | 1 |
| Pathologic Type | Adenoid cystic carcinoma | 1 |
|  | Poorly differentiated carcinoma | 1 |
|  | Salivary duct carcinoma | 1 |
|  | Squamous Cell Carcinoma | 57 |
| HPV Status | HPV Negative | 3 |
|  | HPV Positive | 25 |
|  | Not tested | 32 |
| Primary Tumor Subsite | Oropharyngeal | 46 |
|  | Oral Cavity | 5 |
|  | Nasal Cavity | 1 |
|  | Major Salivary Gland | 2 |
|  | Unknown | 6 |

The swallowing structures were manually contoured on the T2-weighted images by a medical student trained in head and neck organ at risk segmentation and edited as necessary by a head and neck radiation oncologist. The contoured structures include the epiglottis, inferior/medial constrictors, anterior digastric muscle, genioglossus, geniohyoid, bilateral masseters, mylohyoid, soft palate, bilateral lateral pterygoids, bilateral medial pterygoids and oral tongue. The base of tongue and superior constrictor muscles were contoured but excluded from analysis because these structures had GTV involvement in the majority of patients. Additionally, individual structures were excluded for each patient if there was GTV infiltration of that structure.

### Atlas-based Autosegmentation

Atlas-based autosegmentation (ABAS) was performed using the random forest algorithm in ADMIRE (Elekta AB; Stockholm, Sweden). The algorithm first deformably registers each atlas image to the target image and extracts voxel-wise features from the aligned atlases. These features are then used to train a random forest classifier, which predicts the most probable segmentation label for each voxel in the target image based on patterns learned from the atlas data. By integrating information from multiple atlases, this approach reduces errors from individual registrations and improves segmentation accuracy compared to traditional majority voting or intensity-based fusion methods [18]

The 60 HNC patients were split into two groups: 25 patients were used as atlas images for the ABAS model training, and 35 patients were used to test each iteration of the ABAS. Atlas patients were randomly selected from the patients who did not require any structure exclusions due to GTV infiltration, ensuring that all iterations of the ABAS were trained with complete datasets.

To evaluate ABAS performance across different training sizes, autosegmentation was performed using 1–25 atlases (specifically 1–10, 12, 15, 20, 25). For each atlas count, all 35 test patients were autosegmented. A bootstrap resampling approach was used to reduce selection bias and account for variability in atlas selection. For each test case, a random subset of atlases was drawn from the 25-patient training cohort and applied iteratively. The number of bootstrap iterations decreased with increasing atlas count due to reduced variability with larger sample sizes: 25 iterations for 1–2 atlases, 15 for 3, 10 for 4, 5 for 5–12, and 3 for 15 and 20. The 25-atlas configuration used a single iteration, as all training patients were included. An in-house Python script was used to automate the bootstrapping process and interface with ADMIRE for registration and segmentation. Execution time was recorded for each iteration. All ABAS computations were performed on a Microsoft Windows HP ZB G4 Workstation with a Intel(R) Xeon(R) Gold 6132 CPU and NVIDIA Quadro P620 GPU. All data will be shared via Figshare at DOI:10.6084/m9.figshare.29656715.

### Evaluation of Autosegmentation and Statistical Analysis

Dice Similarity Coefficient (DSC), Surface DSC (SDSC), Average Surface Distance (ASD), and Hausdorff 95 (HD95) were calculated with an in-house Python script to evaluate autosegmentation performance against physician-defined ground truth segmentations. Ground-truth test masks were compared to ABAS-generated masks for each region of interest (ROI). Geometric accuracy was assessed using the surface-distances Python package [19] along with custom in-house analysis code, which is publicly available on GitHub (https://github.com/kwahid/ABAS_swallowing_structures).

DSC, SDSC, ASD and HD95 were used to evaluate autosegmentation performance against physician-defined ground truth segmentations [20–22]. The median, range and standard deviation of these metrics were calculated for each ROI and atlas number across all patients and bootstrap iterations (1-12, 15,20,25). The median time for each atlas number was also calculated. Hsu’s multiple comparisons with the best (MCB) analysis was used in JMP version 18 (SAS Institute Inc., Cary, NC) to first determine which atlas number was the best performing for each ROI and comparison metric and then to determine which atlas numbers were statistically similar to the best [23]. The lowest atlas count that was statistically similar to the best performing atlas count was selected as the minimum atlas count without compromising accuracy. A qualitative review of common segmentation failures was then performed.

## Results

### Patient Characteristics

Table 1 provides a summary of the demographic and clinical characteristics of the 60 patients included in this study.

### Segmentation Time by Atlas Count

Figure 1 displays a box plot of segmentation time as a function of atlas count, demonstrating a strong linear increase in computational time (R^2^ = 0.9989) with additional atlases. Median processing time ranged from ∼10 seconds with 1 atlas to ∼950 seconds with 25 atlases. While segmentation time remained low and consistent for 1–5 atlases (median <150 seconds, narrow IQRs), higher atlas counts (≥10) were associated with progressively longer and more variable computation times. Notably, processing times exceeded 750 seconds at 20 atlases and 950 seconds at 25 atlases, with marked increases in variance at the highest counts.

**Figure 1:**
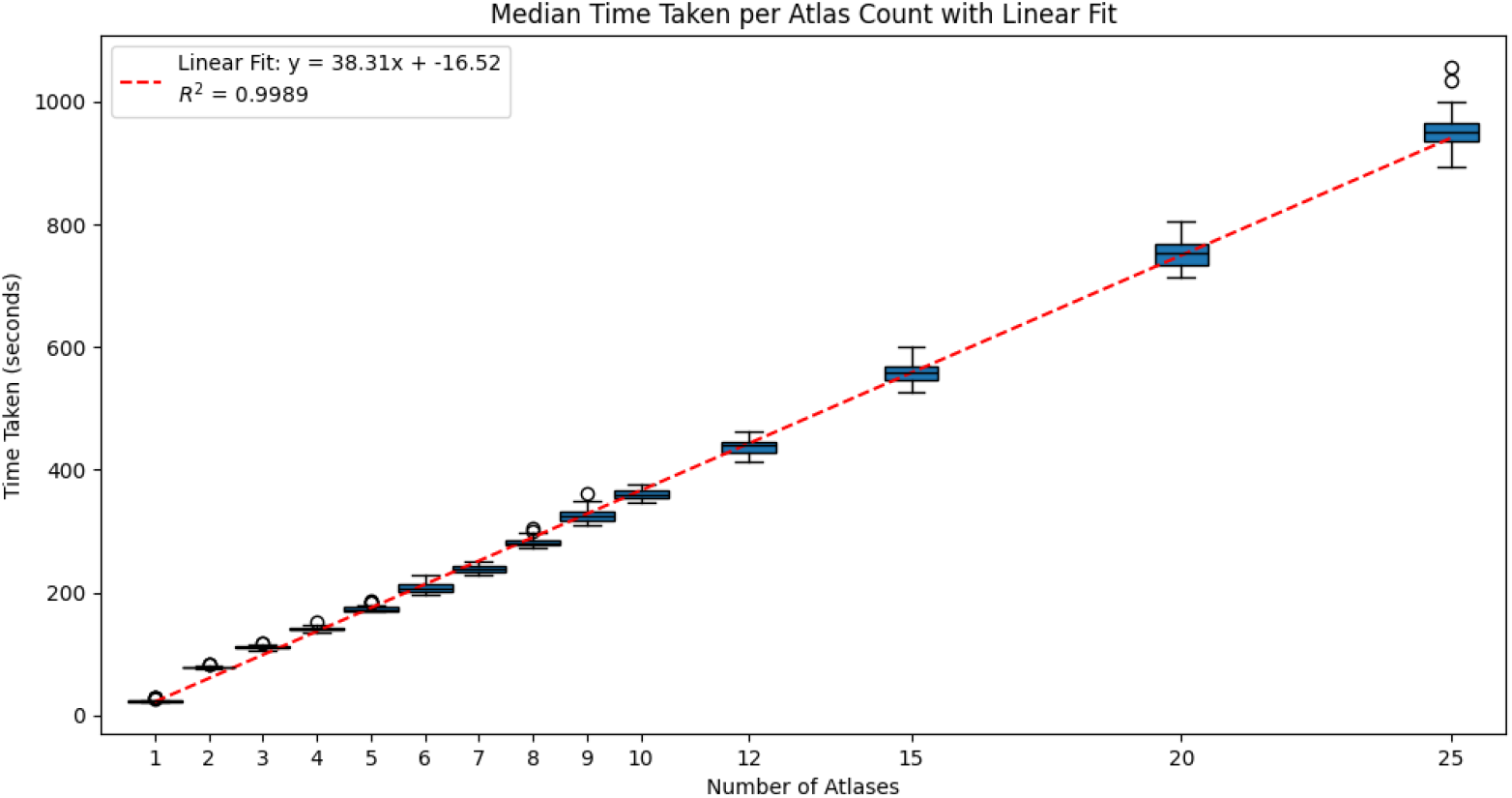
Median segmentation time per atlas count with linear fit. Box plots represent the distribution of processing times at each atlas count, with median values increasing linearly as the number of atlases increases. The red dashed line denotes a linear regression fit (R^2^ = 0.9989).

### Segmentation Accuracy Across Atlas Counts

As shown in Table 2, the fewest atlases required to achieve statistical equivalence with the optimal count ranged from 2– 4 for DSC and 1–4 across the remaining metrics (SDSC, ASD, HD95). Optimal performance was observed with 8–12 atlases for DSC, 8–25 for SDSC, 5–15 for ASD, and 6–25 for HD95.

**Table 2.** Optimal atlas counts for swallowing muscle autosegmentation across geometric accuracy metrics. DSC = Dice Similarity Coefficient, SDSC = Surface Dice Similarity Coefficient, ASD = Average Surface Distance, HD95 = 95th Percentile Hausdorff Distance. Min = Minimum number of atlases required to achieve statistical equivalence with the best-performing atlas count; Best = Atlas count with the highest segmentation performance for each metric.

| ROI | DSC |  | SDSC |  | ASD |  | HD95 |  |
| --- | --- | --- | --- | --- | --- | --- | --- | --- |
|  | Min | Best | Min | Best | Min | Best | Min | Best |
| Epiglottis | 2 | 8 | 1 | 10 | 1 | 15 | 1 | 15 |
| Inferior Pharyngeal Constrictor | 3 | 8 | 2 | 8 | 2 | 7 | 3 | 8 |
| Medial Pharyngeal Constrictor | 2 | 10 | 1 | 9 | 1 | 9 | 1 | 7 |
| Anterior Digastric | 4 | 10 | 4 | 25 | 4 | 12 | 4 | 25 |
| Genioglossus | 2 | 5 | 2 | 15 | 2 | 5 | 2 | 6 |
| Geniohyoid | 2 | 5 | 4 | 20 | 3 | 10 | 3 | 20 |
| Left Masseter | 2 | 7 | 3 | 25 | 3 | 7 | 3 | 20 |
| Right Masseter | 2 | 10 | 2 | 20 | 2 | 7 | 2 | 10 |
| Mylohyoid | 2 | 10 | 3 | 20 | 2 | 10 | 3 | 15 |
| Left Lateral Pterygoid | 2 | 12 | 3 | 12 | 3 | 12 | 3 | 12 |
| Right Lateral Pterygoid | 3 | 12 | 3 | 25 | 3 | 12 | 3 | 25 |
| Left Medial Pterygoid | 2 | 5 | 2 | 10 | 2 | 7 | 2 | 10 |
| Right Medial Pterygoid | 2 | 8 | 3 | 20 | 3 | 15 | 2 | 20 |
| Oral Tongue | 3 | 8 | 2 | 8 | 3 | 8 | 2 | 8 |
| Soft Palate | 2 | 5 | 2 | 25 | 2 | 5 | 2 | 12 |

Figure 2 illustrates DSC performance across atlas counts by structure, with minimum and optimal atlas numbers determined using Hsu’s MCB analysis. Corresponding plots for SDSC, ASD, and HD95 are provided in Supplemental Figures S1–S3. Among cases using ≥2 atlases, all structures except the pharyngeal constrictors achieved median DSC >0.75, SDSC >0.89, and ASD <1 mm. Median HD95 was <4 mm for all structures with ≥2 atlases, except the constrictors. The epiglottis, genioglossus, and mylohyoid demonstrated the highest segmentation accuracy across all metrics (median DSC >0.81, SDSC >0.98, ASD <0.5 mm). The anterior digastric also performed well (median DSC >0.81, ASD <0.4 mm), though median HD95 exceeded 2 mm. The middle and inferior pharyngeal constrictors had the lowest accuracy across all metrics, with median DSC <0.70, median SDSC near the 0.89 threshold, and the highest median ASD and HD95 values among all ROIs. Despite this, all ROIs demonstrated median ASD <1 mm using ≥2 atlases.

**Figure 2:**
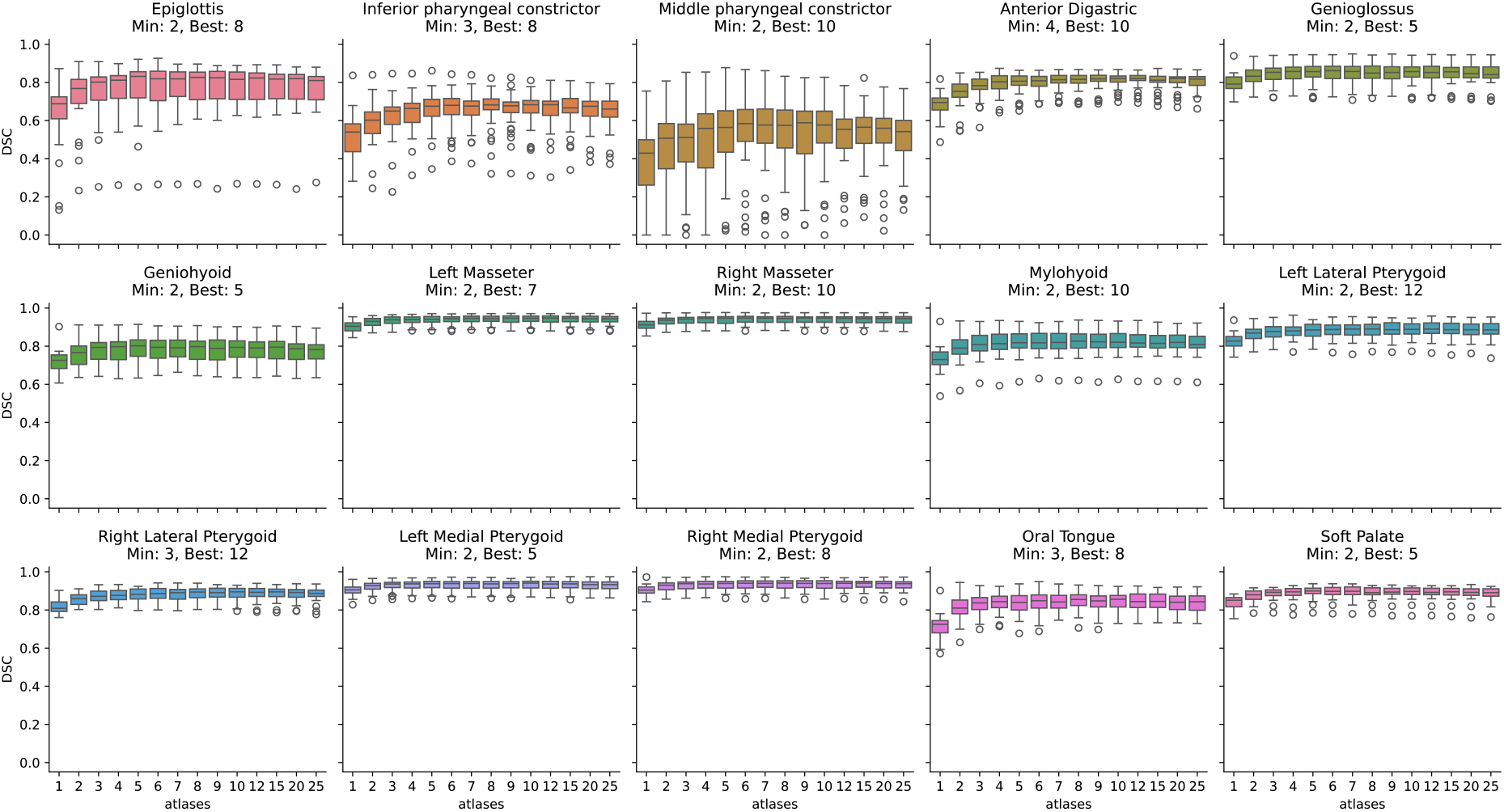
Box plot depicting median and interquartile range of DSC per atlas count. The dashed lines represent the optimal (Best) and minimum (Min) statistically similar atlas number for each structure per Hsu’s analysis with the results summarized in Table 2 for DSC, SDSC, ASD and HD95. The circles represent outliers.

While most structures reached statistical equivalence with as few as 1–3 atlases, the epiglottis and constrictors demonstrated greater variability, with wider interquartile ranges and more frequent outliers at lower atlas counts. Surface-based metrics (ASD and HD95) for the inferior and medial constrictors typically stabilized with 5–7 atlases, while the epiglottis required ≥4. Nevertheless, even these structures achieved statistical equivalence with 1–2 atlases in many cases.

### Outlier Analysis

We performed a qualitative review of segmentation failures across multiple ABAS iterations, focusing on the three structures with the highest number of outlier cases: the epiglottis and the medial and inferior constrictor muscles. A representative failure case for the epiglottis is shown in Figure 3. In this example, a left-sided base of tongue tumor abuts the epiglottis without clear anatomical demarcation, resulting in ABAS-generated contours encroaching into the tumor and compromising segmentation accuracy. This patient demonstrated the poorest epiglottis performance across three of the four evaluated metrics (DSC, SDSC, and ASD) across all atlas counts. Among the broader cohort, many patients had oropharyngeal tumors in close proximity to the epiglottis, a factor that frequently obscured normal anatomical boundaries and likely contributed to the overall reduced segmentation performance for this structure.

**Figure 3:**
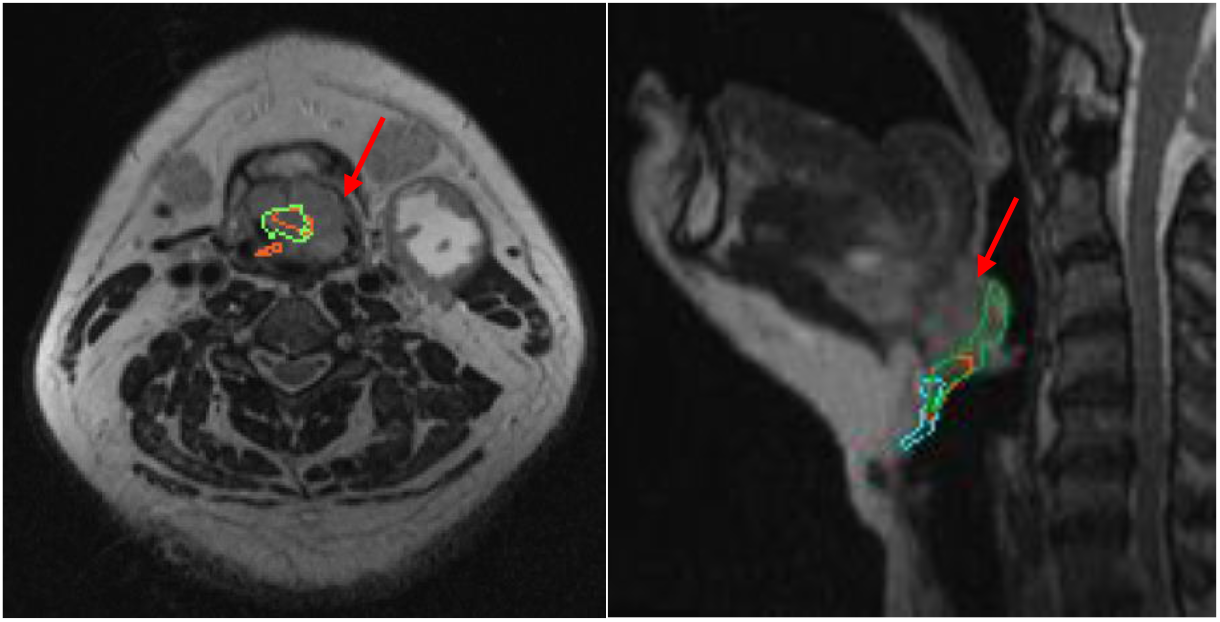
Evaluation of epiglottis contours in a single example patient with a base of tongue tumor (red arrow). The blue contour represents the ground truth. The ABAS contours using 2 atlases and 4 atlases are shown in red and green respectively. A) on the left is an Axial view of T2 MRI with ABAS contours seen within tumor. B) On the right is a sagittal view of T2 MRI the ABAS contours sitting within the tumor the ground truth contour (blue) is appropriately covering the epiglottis inferior to the tumor.

Evaluation of constrictor contours in two separate example patients. A) On the left is a T2 MRI axial view of a patient with a left sided palatine tonsil tumor (outside of view) that shows dropout of the ABAS inferior constrictor contour with the light blue contour representing the ground truth and the green contour representing the ABAS contour using 3 atlases. B) On the right is a T2 MRI sagittal view of a patient with a left sided base of tongue tumor (outside of view) that shows poor delineation between the constrictors at the transition point with the blue and light blue contours representing the ground truth inferior and medial constrictors, respectively, while the green and dark green represent the ABAS contours of the inferior and medial constrictors using 5 atlases.

For the constrictor muscles, two recurring sources of segmentation error were identified: (1) the presence of discontinuities or contour dropout in the ABAS-generated contours where the ground truth contours thinned but remained anatomically continuous, and (2) inconsistency in the delineation between the medial and inferior constrictor segments at their transition point. These issues likely contributed to degraded performance, particularly in the two worst-performing patients shown in Figures 4A and 4B.

**Figure 4:**
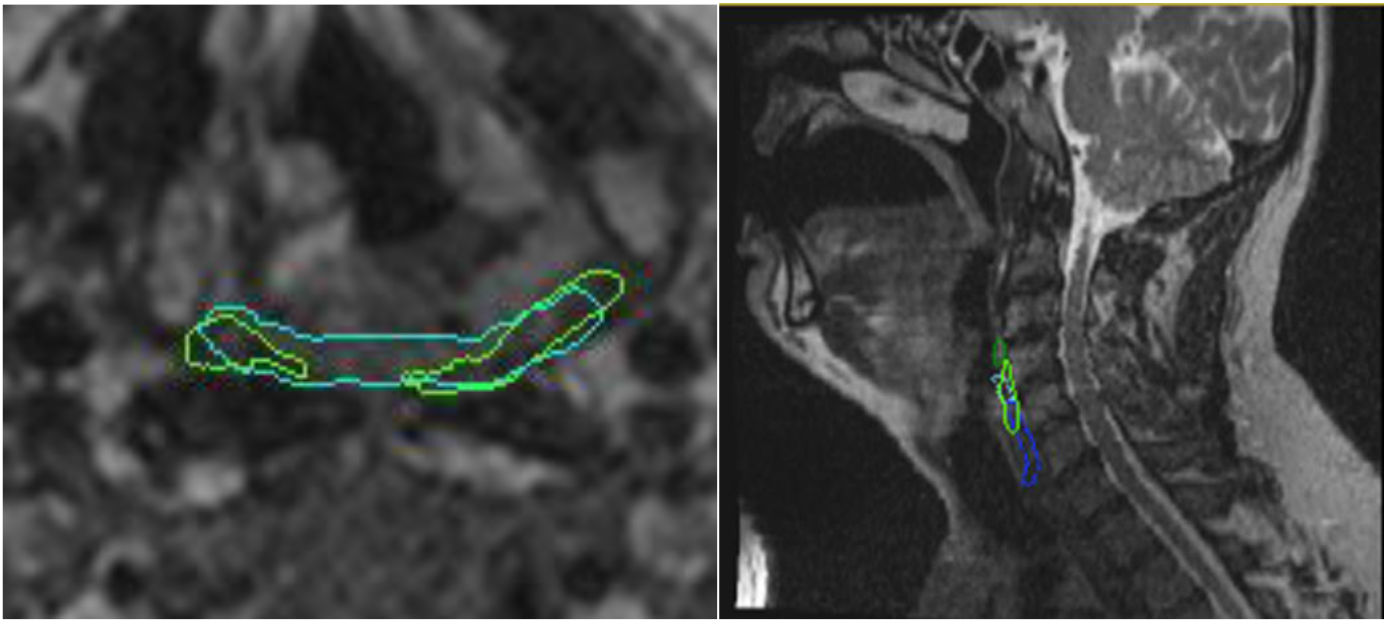
Evaluation of constrictor contours in two separate example patients. A) On the left is a T2 MRI axial view of a patient with a left sided palatine tonsil tumor (outside of view) that shows dropout of the ABAS inferior constrictor contour with the light blue contour representing the ground truth and the green contour representing the ABAS contour using 3 atlases. B) On the right is a T2 MRI sagittal view of a patient with a left sided base of tongue tumor (outside of view) that shows poor delineation between the constrictors at the transition point with the blue and light blue contours representing the ground truth inferior and medial constrictors, respectively, while the green and dark green represent the ABAS contours of the inferior and medial constrictors using 5 atlases.

## Discussion

This study demonstrates that atlas-based autosegmentation (ABAS) using T2-weighted MRI achieves accurate segmentation of swallowing muscles with as few as 2–4 atlases, significantly reducing computational time while maintaining geometric accuracy. Across all metrics—DSC, SDSC, ASD, and HD95—the minimum number of atlases required for statistical equivalence ranged from 1 to 4, while the best-performing atlas count varied from 5 to 25, depending on the structure and metric. DSC and SDSC followed similar trends, with most structures achieving peak performance between 8 and 12 atlases. However, some muscles, including the digastric anterior, masseters, and pterygoids, required up to 25 atlases for optimal SDSC performance, reflecting increased segmentation variability. ASD and HD95 exhibited greater variability, with higher best-performing atlas counts in the epiglottis, digastric anterior, and pterygoid muscles, where 12 to 25 atlases were necessary to achieve optimal geometric accuracy. The impact of segmentation time was also notable, with computation times increasing from approximately 10 seconds for 1 atlas to over 950 seconds for 25 atlases. These findings suggest that while low atlas counts (2–4) are sufficient for many structures, some require additional atlases for consistency, though at the cost of significantly longer processing times and thus possibly impacting clinical workflows.

These findings emphasize the importance of balancing segmentation accuracy with computational efficiency, particularly in high-throughput radiotherapy planning environments. While larger atlas sets improve delineation of certain complex structures, the substantial increase in processing time presents a practical limitation [24]. Optimizing atlas selection and exploring alternative autosegmentation strategies, such as deep learning–based approaches, could enhance segmentation consistency while maintaining feasible processing times [25]. Future work should assess whether hybrid atlas-based and deep learning models can further improve segmentation performance, particularly for anatomically complex swallowing muscles prone to greater variability.

The failure modes shown in Figures 3 and 4 underscore the limitation of ABAS in anatomically ambiguous regions or in the presence of adjacent tumor volumes. Such errors are consistent with prior reports demonstrating that atlas or AI-based segmentation approaches often struggle when anatomical boundaries are distorted by tumor proximity or by anatomical variation not seen in the training images [26]. Thin, elongated structures like the digastric muscles are prone to segmentation discontinuities due to poor soft-tissue contrast and the risk of contour dropout [27], leading to holes in contours which were observed in our study. Additionally, prior studies have shown that transitions between adjacent muscle groups, such as the medial and inferior constrictors, exhibit high inter-observer variability [28]. Although our ground truth contours were generated by a single expert, the inherent ambiguity in defining these transitions likely contributed to inconsistent ABAS performance in these regions [29]. These findings suggest that future models may benefit from incorporating anatomical continuity constraints and tumor-aware segmentation strategies. While these geometric failures are apparent, further analysis is needed to determine if these failures are clinically relevant from a dosimetric standpoint, as prior studies point out that poor geometric performance may not necessarily result in poor dosimetry [27].

While our study highlights the sample efficiency of atlas-based autosegmentation (ABAS), it is important to acknowledge that recent work suggests U-Net–based deep learning (DL) models can also achieve strong performance with relatively modest training cohorts. These models, particularly U-Net architectures, have demonstrated robust performance across medical image segmentation tasks even under data-constrained conditions [30,31]. These findings suggest that while ABAS may retain advantages in extremely low-data settings (e.g., 1–2 patients), DL-based approaches remain highly viable alternatives and may offer improved scalability and generalizability as datasets grow.

This study has several limitations. First, it was conducted using a single atlas-based autosegmentation platform (ADMIRE, Elekta), and results may not generalize to other commercial or open-source ABAS systems. Second, all MRI data were acquired at a single institution using standardized simulation protocols, which may not reflect the variability encountered across institutions in terms of image quality, acquisition parameters, or magnetic field strength. Third, the ground truth segmentations were generated by a trained medical student with expert review using standardized protocols but without assessment of inter-observer variability, which may affect the generalizability of the ground truth reference standard. Finally, we did not explicitly account for the impact of tumor location or volume on autosegmentation performance, though qualitative review of our outliers suggests that tumor proximity did play a critical role in failure modes. Future studies should consider incorporating tumor-aware modeling and validating these findings in multi-institutional datasets.

Ultimately, the ability to efficiently and accurately segment swallowing muscles on MRI has direct clinical implications. These structures play a critical role in post-treatment swallowing function, and retrospective studies have demonstrated correlations between dose to individual swallowing muscles and dysphagia risk [3,6,32]. By enabling consistent and reproducible segmentation across patients, ABAS provides a practical foundation for evaluating dose-response relationships and informing NTCP modeling [33]. These models may inform individualized dose constraints and support treatment planning strategies to reduce dysphagia risk as MRI-guided radiotherapy becomes more widely adopted in head and neck cancer.

## Conclusion

To our knowledge, this is the first study to systematically evaluate MRI-based atlas autosegmentation for dysphagia-relevant swallowing muscles in head and neck cancer. By assessing segmentation accuracy, processing time, and patterns of failure across atlas sizes, we identify practical tradeoffs that directly impact clinical workflows, particularly in adaptive or high-throughput radiotherapy settings. As MRI becomes more integrated into radiotherapy planning and OAR-sparing remains a clinical priority, these findings provide timely insight and can inform the development of hybrid or AI-driven segmentation strategies to support efficient, anatomically accurate treatment planning.

## Data Availability

All data produced in the present study are available upon reasonable request to the authors

https://github.com/kwahid/ABAS_swallowing_structures

## Funding Acknowledgement

KW and BAM received funding support from the National Institutes of Health (NIH) National Cancer Institute (NCI) for this project under the Image Guided Cancer Therapy Training Program (T32CA261856). SS was supported during project execution by the Swiss Cancer League Bursary (BIL KLS-4300-08-2017)SYL. SYL and CDF received related funding for data collection from NIH National Institute of Dental and Craniofacial Research (NIDCR; R56DE02524, R01DE02524, R01DE028290); SYL, KAH, and CDF received related technical development support from NCI (R01CA218148). CDF received relevant funding for data analysis from a joint National Science Foundation (NSF)/NCI award through NCI (R01CA257814). Infrastructure support was provided by the MD Anderson Cancer Center Core Support Grant (CCSG) Image-Guided Interventions and Insights (III) Program (P30CA016672).

## Data sharing statement

In accordance with NOT-OD-21-013, Final NIH Policy for Data Management and Sharing, anonymized/de-identified data that support the findings of this study are openly available in an NIH-supported generalist scientific data repository (figshare) at http://doi.org/XXXXXX no later than the time of an associated publication.

## Public access policy compliance

In accordance with NOT-OD-25-049, Supplemental Guidance to the 2024 NIH Public Access Policy: Government Use License and Rights,: “This manuscript is the result of funding in whole or in part by the National Institutes of Health (NIH). It is subject to the NIH Public Access Policy. Through acceptance of this federal funding, NIH has been given a right to make this manuscript publicly available in PubMed Central upon the Official Date of Publication, as defined by NIH.”

## Pre-print Statement

Consistent with NOT-OD-17-050, Reporting Preprints and Other Interim Research Products, as “NIH encourages investigators to use interim research products, such as preprints, to speed the dissemination and enhance the rigor of their work”, a pre-peer reviewed deposition of the initial submission version of the manuscript has been deposited for public access ([insert DOI for medrxiv deposition]).

## CRediT statement

In accordance with the Contributor Roles Taxonomy (CRediT, https://credit.niso.org/), the contributing authors have designated responsibilities and individual author attribution. The corresponding author(s) (KB) assume(s) responsibility for role assignment, and all contributors have been given the opportunity to review and confirm assigned roles.

- ZB: Methodology, Formal analysis, Investigation, Data curation, Visualization, Writing, Review & Editing
- KW: Methodology, Software, Validation, Formal analysis, Writing, Review & Editing
- SS: Data curation, Investigation, Review & Editing
- RD: Data curation, Review & Editing
- CSS: Investigation, Data curation, Review & Editing
- KAH: Conceptualization, Review & Editing
- SYL: Resources, Funding acquisition, Review & Editing
- CDF: Conceptualization, Funding acquisition, Supervision, Project administration, Writing, Review & Editing
- BAM: Conceptualization, Methodology, Supervision, Project administration, Writing, Review & Editing, Corresponding Author

## References

1. De Felice F, de Vincentiis M, Luzzi V, et al. Late radiation-associated dysphagia in head and neck cancer patients: Evidence, research and management. Oral Oncol. 2018;77:125–30. doi:10.1016/j.oraloncology.2017.12.021.

2. Caudell JJ, Schaner PE, Desmond RA, et al. Dosimetric factors associated with long-term dysphagia after definitive radiotherapy for squamous cell carcinoma of the head and neck. Int J Radiat Oncol Biol Phys. 2010;76(2):403–9. doi:10.1016/j.ijrobp.2009.02.017.

3. Dirix P, Abbeel S, Vanstraelen B, et al. Dysphagia after chemoradiotherapy for head-and-neck squamous cell carcinoma: Dose-effect relationships for the swallowing structures. Int J Radiat Oncol Biol Phys. 2009;75(2):385– 92. doi:10.1016/j.ijrobp.2008.11.041.

4. Jensen K, Lambertsen K, Grau C. Late swallowing dysfunction and dysphagia after radiotherapy for pharynx cancer: Frequency, intensity and correlation with dose and volume parameters. Radiother Oncol. 2007;85(1):74– 82. doi:10.1016/j.radonc.2007.06.004.

5. Hutcheson KA, Lewin JS, Barringer DA, et al. Late dysphagia after radiotherapy-based treatment of head and neck cancer. Cancer. 2012;118(23):5793–9. doi:10.1002/cncr.27631.

6. Eisbruch A, Kim HM, Feng FY, et al. Chemo-IMRT of oropharyngeal cancer aiming to reduce dysphagia: Swallowing organs late complication probabilities and dosimetric correlates. Int J Radiat Oncol Biol Phys. 2011;81(3):e93–9. doi:10.1016/j.ijrobp.2010.12.067.

7. Schmidt MA, Payne GS. Radiotherapy planning using MRI. Phys Med Biol. 2015;60(22):R323–61. doi:10.1088/0031-9155/60/22/R323.

8. Cardenas CE, Mohamed ASR, Yang J, et al. Head and neck cancer patient images for determining auto-segmentation accuracy in T2-weighted magnetic resonance imaging through expert manual segmentations. Med Phys. 2020;47(5):2317–22. doi:10.1002/mp.13942.

9. Kosmin M, Ledsam J, Romera-Paredes B, et al. Rapid advances in auto-segmentation of organs at risk and target volumes in head and neck cancer. Radiother Oncol. 2019;135:130–40. doi:10.1016/j.radonc.2019.03.004.

10. Van de Velde J, Wouters J, Vercauteren T, et al. Optimal number of atlases and label fusion for automatic multiatlas-based brachial plexus contouring in radiotherapy treatment planning. Radiat Oncol. 2016;11:1. doi:10.1186/s13014-015-0579-1.

11. Greenham S, Dean J, Fu CK, et al. Evaluation of atlas-based auto-segmentation software in prostate cancer patients. J Med Radiat Sci. 2014;61(3):151–8. doi:10.1002/jmrs.64.

12. Aljabar P, Heckemann RA, Hammers A, et al. Multi-atlas based segmentation of brain images: Atlas selection and its effect on accuracy. Neuroimage. 2009;46(3):726–38. doi:10.1016/j.neuroimage.2009.02.018.

13. Anders LC, Stieler F, Siebenlist K, et al. Performance of an atlas-based autosegmentation software for delineation of target volumes for radiotherapy of breast and anorectal cancer. Radiother Oncol. 2012;102(1):68– 73. doi:10.1016/j.radonc.2011.08.043.

14. Han X, Hoogeman MS, Levendag PC, et al. Atlas-based auto-segmentation of head and neck CT images. Med Image Comput Comput Assist Interv. 2008;11(Pt 2):434–41. doi:10.1007/978-3-540-85990-1_52.

15. Breiman L. Random forests. Mach Learn. 2001;45(1):5–32.

16. Han X. WE-E-213CD-06: A locally adaptive, intensity-based label fusion method for multi-atlas auto-segmentation. Med Phys. 2012;39(6Part27):3960. doi:10.1118/1.4736162.

17. de Vette SPM, van Rijn-Dekker MI, Van den Bosch L, Keijzer K, Neh H, Chu H, et al. Evaluation of a comprehensive set of normal-tissue complication-probability models for patients with head and neck cancer in an international cohort. Oral Oncol. 2025;163:107224. doi:10.1016/j.oraloncology.2025.107224

18. McDonald BA, Ho TY, Ahunbay E, et al. Investigation of autosegmentation techniques on T2-weighted MRI for off-line dose reconstruction in MR-linac workflow for head and neck cancers. Med Phys. 2024;51(4):2332–44. doi:10.1002/mp.16582.

19. Park B, Choi J, Kim S, et al. Development and validation of a deep learning–based patient decision aid for prostate cancer radiotherapy. J Med Internet Res. 2021;23(5):e26151. doi:10.2196/26151.

20. Yeghiazaryan V, Voiculescu I. Family of boundary overlap metrics for the evaluation of medical image segmentation. J Med Imaging (Bellingham). 2018;5(1):015006. doi:10.1117/1.JMI.5.1.015006.

21. Brouwer CL, Steenbakkers RJHM, van den Heuvel E, et al. Metrics to evaluate the performance of auto-segmentation for radiation treatment planning: A critical review. Radiother Oncol. 2021;160:185–91. doi:10.1016/j.radonc.2021.05.003.

22. Sharp GC, Fritscher KD, Pekar V, et al. Vision 20/20: Perspectives on automated image segmentation for radiotherapy. Med Phys. 2014;41(5):050902. doi:10.1118/1.4871620.

23. Artman WJ, Nahum-Shani I, Wu T, et al. Power analysis in a SMART design: Sample size estimation for determining the best embedded dynamic treatment regime. Biostatistics. 2020;21(3):432–48. doi:10.1093/biostatistics/kxy064.

24. Aoyama T, Shimizu H, Kitagawa T, et al. Comparison of atlas-based auto-segmentation accuracy for radiotherapy in prostate cancer. Phys Imaging Radiat Oncol. 2021;19:126–30. doi:10.1016/j.phro.2021.08.002.

25. Matoska T, Patel M, Liu H, et al. Review of deep learning-based autosegmentation for clinical target volume: Current status and future directions. Adv Radiat Oncol. 2024;9(5):101470. doi:10.1016/j.adro.2024.101470.

26. Vrtovec T, Močnik D, Strojan P, et al. Auto-segmentation of organs at risk for head and neck radiotherapy planning: From atlas-based to deep learning methods. Med Phys. 2020;47(9):e929–50. doi:10.1002/mp.14320.

27. Petkar I, Rooney K, Roe JW, et al. Inter-observer variation in delineating the pharyngeal constrictor muscle as organ at risk in radiotherapy for head and neck cancer. Front Oncol. 2021;11:644767. doi:10.3389/fonc.2021.644767.

28. Pérez-Ortiz M, García-Figueiras R, Pardo-Montero J. Interobserver and intermodality variability analysis in OAR contouring from head and neck CT and MR images. Med Phys. 2023;50(6):3660–71. doi:10.1002/mp.16924.

29. Amjad A, Xu J, Thill D, et al. General and custom deep learning autosegmentation models for organs in head and neck, abdomen, and male pelvis. Med Phys. 2022;49(3):1686–700. doi:10.1002/mp.15507.

30. Tappeiner E, Pröll S, Fritscher K, et al. Training of head and neck deep learning autosegmentation models with limited data: A study on the impact of training sample size. Phys Med Biol. 2022;67(19):195002. doi:10.1088/1361-6560/ac2206.

31. Kugelman J, Allman D, Mahajan S, et al. A comparison of deep learning U-Net architectures for posterior segment OCT retinal layer segmentation. Sci Rep. 2022;12(1):14398. doi:10.1038/s41598-022-18646-2.

32. Christianen ME, Langendijk JA, Westerlaan HE, et al. Delineation of organs at risk involved in swallowing for radiotherapy treatment planning. Radiother Oncol. 2011;101(3):394–402. doi: 10.1016/j.radonc.2011.05.015

33. van der Laan HP, van den Bosch S, van der Schaaf A, et al. Swallowing-sparing intensity-modulated radiotherapy (SW-IMRT) in head and neck cancer: Clinical validation of a model-based approach. Radiother Oncol. 2024;190:110072. doi: 10.1016/j.radonc.2023.110044

