## Supplemental Tables 1-3 for "Optimizing Atlas Counts for MRI-Guided Atlas-Based Autosegmentation of Swallowing Muscles in Head and Neck Radiotherapy"

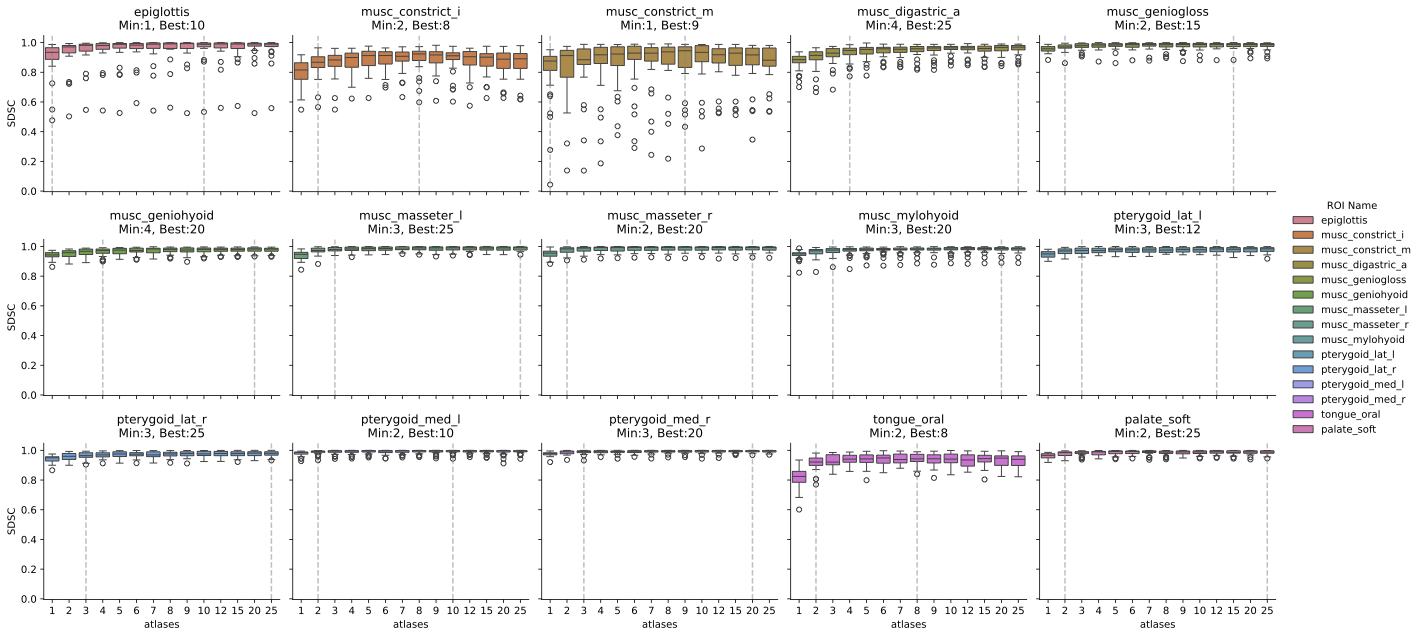

Supplemental Figure 1: Box plot depicting median and interquartile range of SDSC per atlas count. The dashed lines represent the optimal (Best) and minimum (Min) statistically similar atlas number for each structure Per Hsu’s analysis with the results summarized in Table 2 for DSC, SDSC, ASD and HD95. The circles represent outliers.

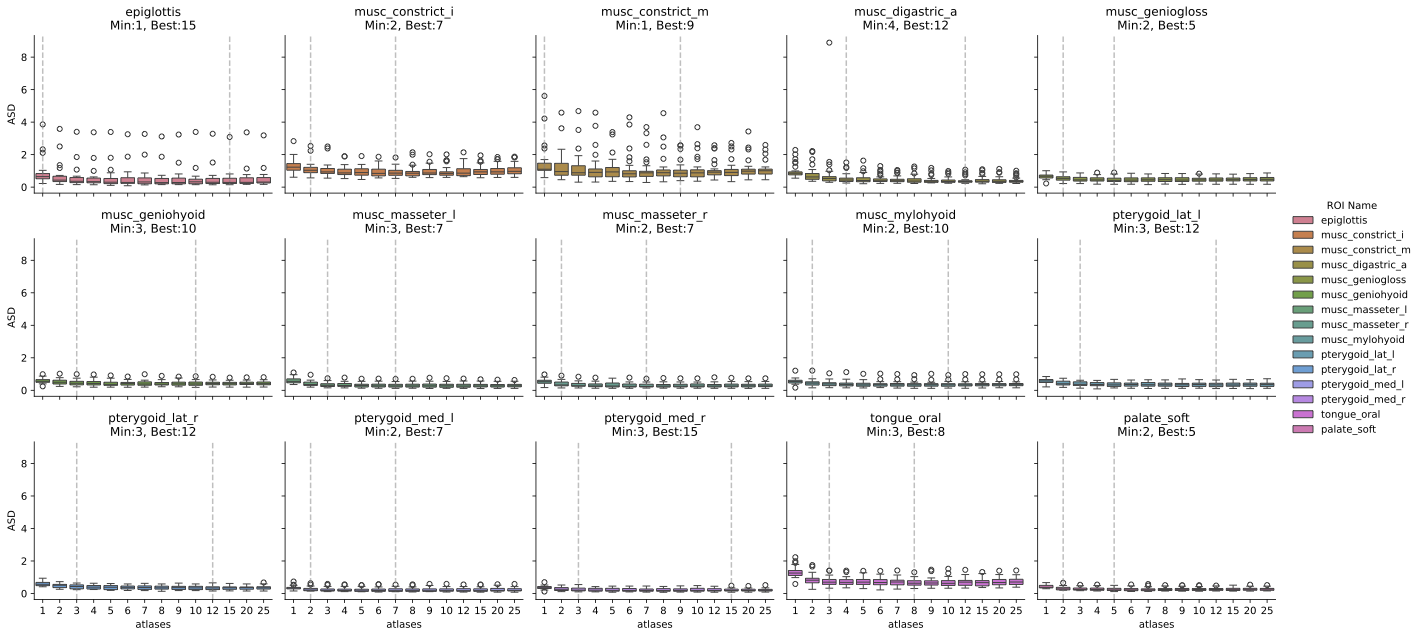

Supplemental Figure 2: Box plot depicting median and interquartile range of ASD per atlas count. The dashed lines represent the optimal (Best) and minimum (Min) statistically similar atlas number for each structure Per Hsu’s analysis with the results summarized in Table 2 for DSC, SDSC, ASD and HD95. The circles represent outliers.

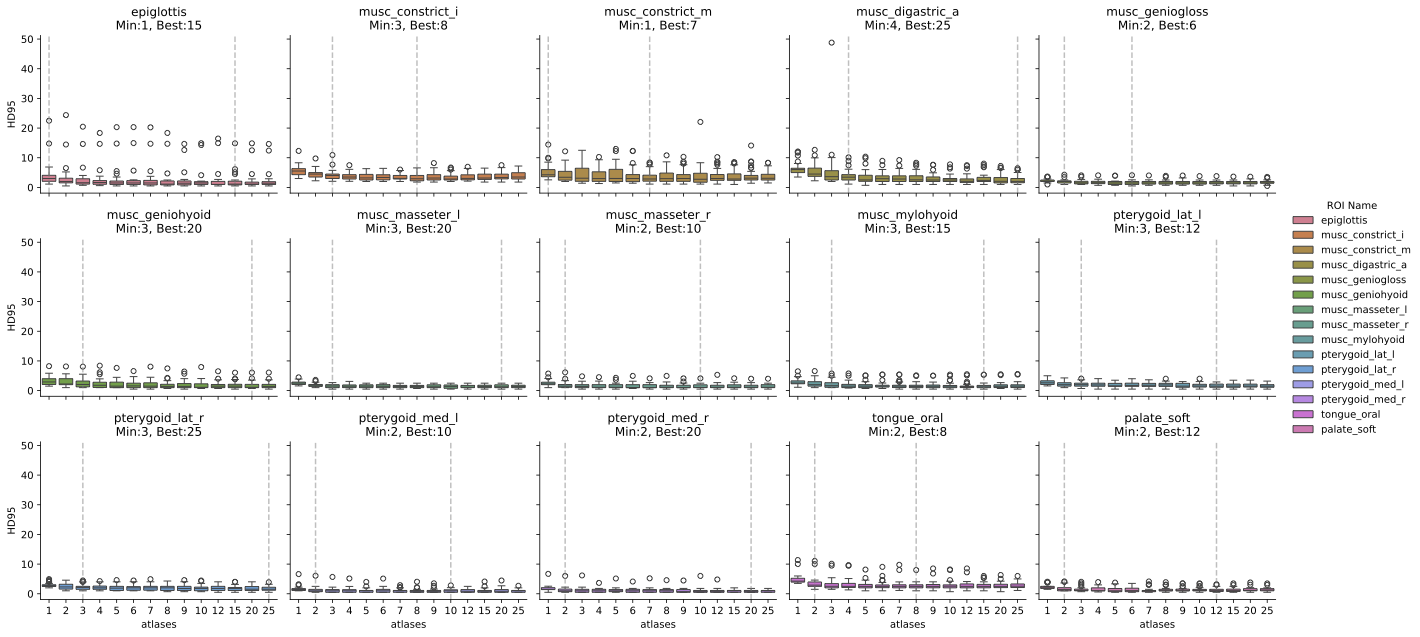

Supplemental Figure 3: Box plot depicting median and interquartile range of HD95 per atlas count. The dashed lines represent the optimal (Best) and minimum (Min) statistically similar atlas number for each structure Per Hsu’s analysis with the results summarized in Table 2 for DSC, SDSC, ASD and HD95. The circles represent outliers.
